# Viable circulating endothelial cells and their progenitors are increased in Covid-19 patients

**DOI:** 10.1101/2020.04.29.20085878

**Authors:** Patrizia Mancuso, Antonio Gidaro, Giuliana Gregato, Alessandro Raveane, Paola Cremonesi, Jessica Quarna, Sonia Caccia, Luca Gusso, Chiara Cogliati, Francesco Bertolini

## Abstract

During the course of Covid-19, the disease caused by the new Coronavirus SARS-CoV-2, thrombotic phenomena and/or diffuse vascular damage are frequent, and viral elements have been observed within endothelial cells. Circulating endothelial cells (CECs) and their progenitors (CEPs) are increased in cardiovascular, thrombotic, infectious and cancer diseases. Using a validated flow cytometry procedure, we found that viable CEPs/mL were significantly increased in Covid-19 patients compared to healthy controls. This increase was observed in patients with mild symptoms and not further augmented in patients with severe symptoms. In patients who recovered, CEPs decreased, but were in a range still significantly higher than normal controls. Regarding mature CECs, in Covid-19 patients their absolute number was similar to those observed in healthy controls, but the viable/apoptotic CEC ratio was significantly different. Both mild and severe Covid-19 patients had significantly more viable CECs compared to healthy controls. Patients who recovered had significantly less CECs/mL when compared to controls as well as to mild and severe Covid-19 patients. A positive correlation was found between the copies of SARS-CoV-2 RNA in the cellular fraction and apoptotic CEPs/mL in severe Covid-19 patients. These findings suggest that CECs and CEPs might be investigated as candidate biomarkers of endothelial damage in Covid-19 patients.

## Introduction

The virus causing the 2020 coronavirus epidemic has been called “severe acute respiratory syndrome coronavirus 2” (SARS-CoV-2). The disease caused by the new Coronavirus has been called Covid-19 (1). During the course of the disease, and particularly in the advanced phase, thrombotic phenomena and/or diffuse vascular damage are frequently observed. Moreover, viral elements have been observed within endothelial cells in Covid-19 patients (2–3). An early assessment of these phenomena is believed to potentially help in selecting patients to be treated more quickly.

Circulating endothelial cells (CECs) and their progenitor counterparts (CEPs) are endothelial cells present in the peripheral blood of healthy subjects (where apoptotic CECs deriving from the turnover of the vascular endothelium are observed) and increased in cardiovascular, thrombotic, infectious and cancer diseases (4–5). A validated and reproducible procedure (6) allows to quantify CECs and CEPs by multiparametric flow cytometry and to divide them into their viable and apoptotic fractions.

In the present work we measured CECs and CEPs in active Covid-19 patients, subjects who had Covid-19, recovered and tested negative in the previous week, and age- and gender-matched healthy controls. SARS-CoV-2-RNA viremia was determined in the plasma and cellular fraction of every Covid-19 patient with digital droplet PCR, an assay known to reduce the frequency of false negatives when comparted to RT-PCR (7).

## Patients and Methods

### Patients and controls

During April 2020 we investigated 22 consecutive active Covid-19 patients (10 severe and 12 mild according to RCP values), and 8 subjects who had Covid-19, recovered and tested negative in the previous week. There were 13 females and 17 males, median age was 52 (range 27–81). Eight age- and gender-matched healthy controls were also investigated.

Patient clinical and laboratory data were retrieved from the electronic clinical record. Laboratory data included whole blood cell count, RCP, D-Dimer, creatinine, IL-6, and ferritin.

### Flow cytometry

CECs and CEPs were evaluated by flow cytometry according to Mancuso et al (6) with little modifications. In brief, blood was collected in EDTA tubes as anticoagulant. After lysis of 7ml of blood with ammonium chloride (NH4Cl), cell were incubated with monoclonal antibodies for 15 minute, at 4°C, in the dark. CECs and CEPs were evaluated by ten-color flow cytometry (Navios EX, Beckman Coulter) using the nuclear staining Syto16 for DNA (Thermo Fisher Scientific, Eisai, Medipost – US), 7-AAD (BD) and a panel of monoclonal antibodies including anti-CD45 (to exclude hematopoietic cells), anti CD34, anti-CD31, (both from Beckman Coulter) and anti-CD146 (BD) as endothelial cell markers. After acquisition of at least 3×10^6^ events, an appropriate gating strategy was used to enumerate viable and apoptotic CECs and CEPs (Kaluza sotware, Beckman Coulter). CECs were identified as DNA+, CD45-, CD31+, CD34+, CD146+ and CEPs as DNA+, CD45-, CD31+, CD34+, CD146-. Viable and Apotptotic cells were identified by 7-AAD detection. The absolute number of the cells was calculated by a dual-platform counting technique using the total number of leukocytes in peripheral blood obtained by hemocytometer, according to routine methods.

### Digital droplet PCR

RNA was extracted from 1200 ml of plasma and 600 ml of packed cellular fraction by QIAamp circulating nucleic acid kit (Qiagen,Germany) and tested by SARS-CoV-2 RNA digital droplet polymerase chain reaction (QX200, Bio-Rad) with CDC primers and probes (Integrated DNA technologies, 2019 nCOV-KIT).

### Ethical approval

The “Studio virologico, immunologico, clinico e genetico sui pazienti con infezione da nuovo coronavirus SARS-CoV-2” was approved with the registration number 2020/ST/049 at the “Sacco” Hospital in Milan.

### Statistical analyses

Normal distribution of the data was assessed using Shapiro Wilk’s normality test and the homogeneity of the variance was checked with the F-test for single comparison while using Bartelett’s test for pairwise comparisons. Single and pairwise comparisons were performed using either the Student’s t-test or the Wilcoxon’s Rank Sum test according on normal or not normal distributions respectively; eventually the pairwise comparison p-values were adjusted by the Benjamini-Hochberg method.

For the correlations, Pearson or Spearman tests were employed based on normal or not normal distributions assessed again using Shapiro Wilk’s normality test

Only significant differences (p < 0.05) were plotted. Statistical analyses and plots were carried out using ggpubr and ggplot packages on R. The codes for these analyses are available upon request.

## Results

As shown in Fig. 1A-G, viable CEPs/mL were significantly increased in Covid-19 patients compared to healthy controls (p<0.001). This increase was larger than those usually observed in other neoplastic or vascular diseases (4–6). Interestingly, the increase was observed in patients with mild symptoms and not further augmented in patients with severe symptoms. Patients who recovered had significantly less CEPs/mL, but in a range still significantly higher than normal controls (p<0.01).

**Fig. 1:**
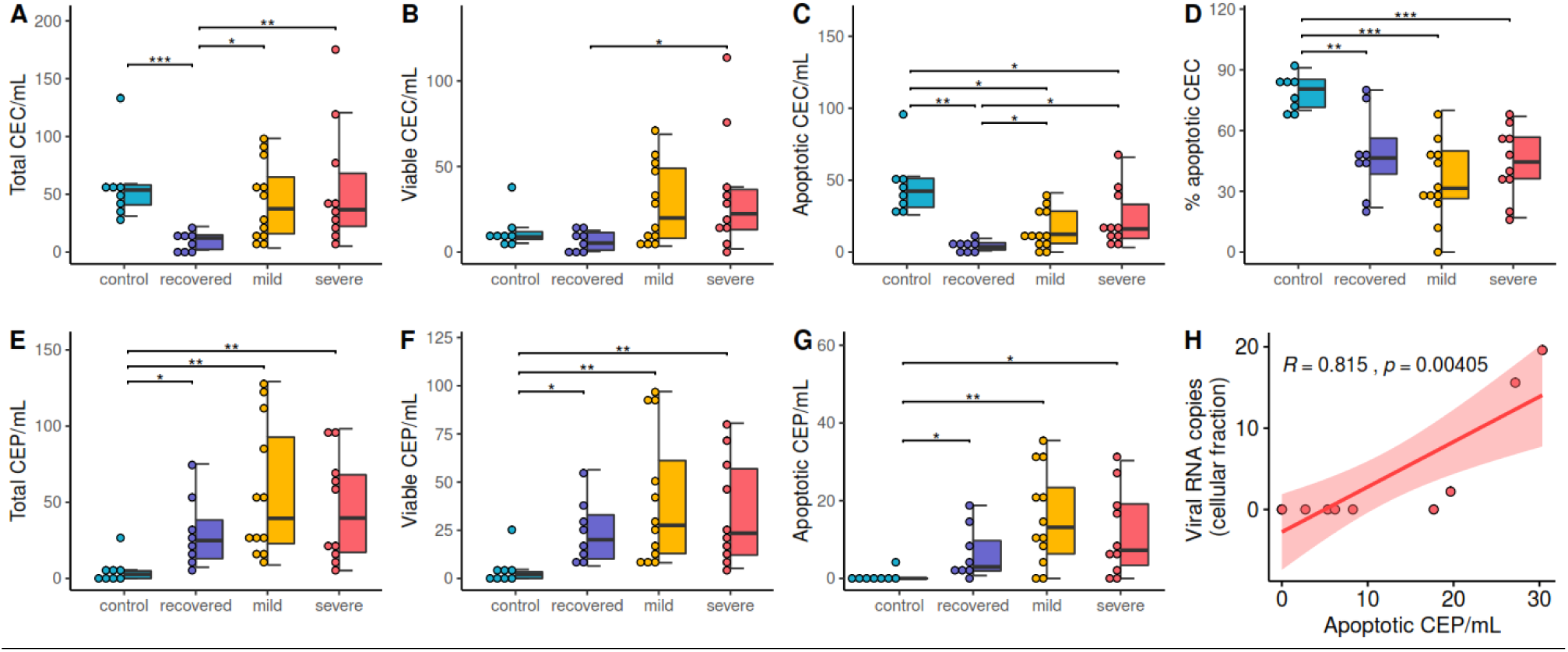
Features of CEP and CEC in Covid-19 patients. A-G) Distributions of CEP and CEC values in the patient groups (* p-value < 0.05, ** p-value < 0.01, *** p-value < 0.001). H) Significative correlation of apoptotic CEP and viral RNA copies in the cellular fraction.

Regarding mature CECs, in Covid-19 patients their absolute number was similar to those observed in healthy controls, but the viable/apoptotic CEC ratio was significantly different (p<0.001). Again, this difference was larger than those usually observed in other neoplastic or vascular diseases (4–6). Patients who recovered had significantly less CECs/mL when compared to controls as well as to mild and severe Covid-19 patients (p<0.01).

SARS-CoV-2 RNA was found in the plasma and/or in the cellular fraction of 6 out of 10 severe Covid-19 patients (60%), and of 2 out of 12 mild Covid-19 patients (1.6%, p=0.353 by X square). An interesting positive correlation (R=0.815, p=0.004) was found between the copies of SARS-CoV-2 RNA in the cellular fraction and apoptotic CEPs/mL in severe Covid-19 patients (Fig. 1H). As shown in Fig. 2, in severe Covid-19 patients other significant correlations were found between apoptotic CEPs and ferritin, total leukocyte count, lymphocytes, monocytes, and basophils. In this subgroup of patients, viable and/or apoptotic CECs were found to correlate with RCP at enrollment and/or at the time of test.

**Fig. 2:**
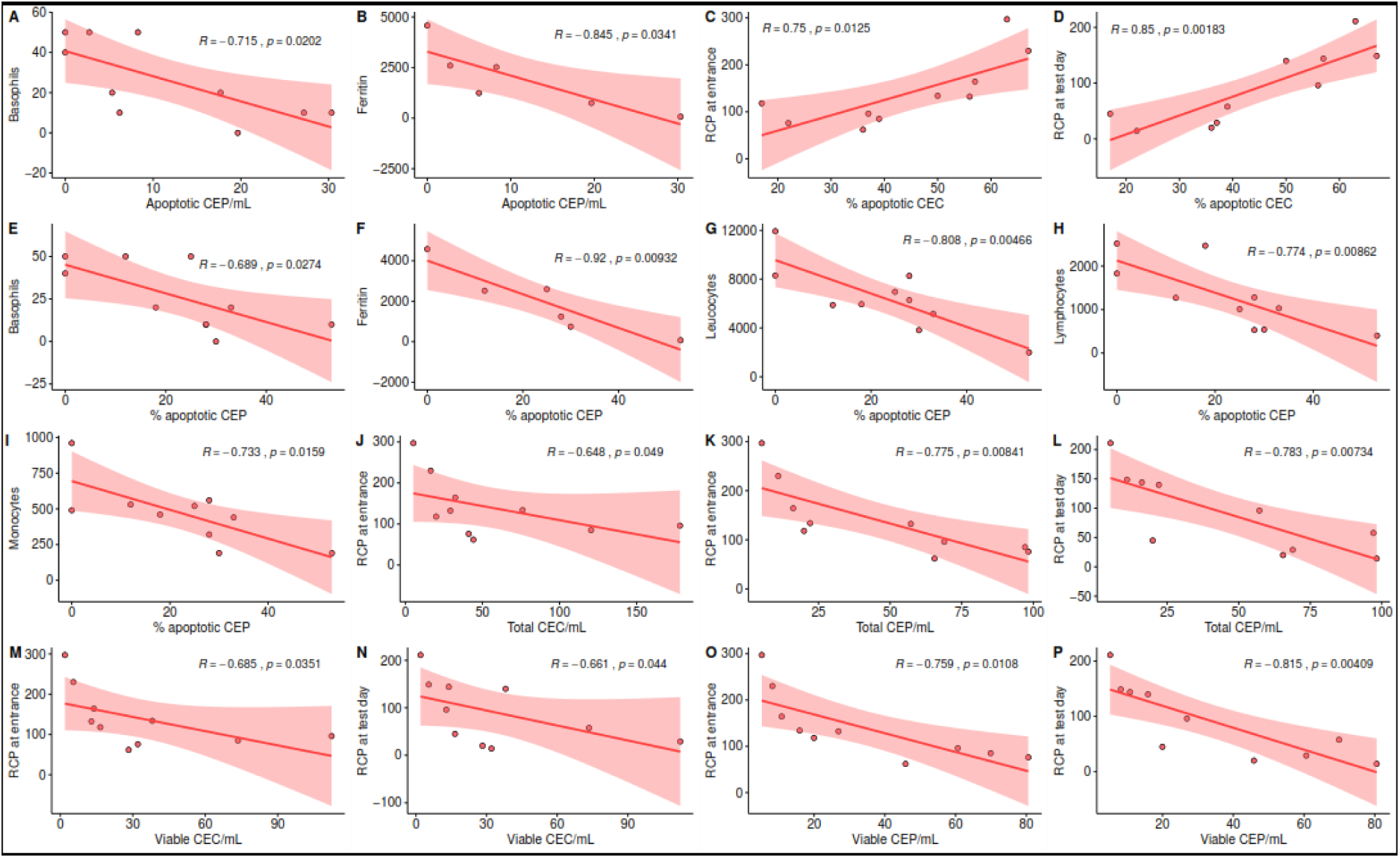
Significant correlations among CEP and CEP categories and hematological parameters in severe Covid-19 patients.

As shown in Fig. 3, in patients affected by mild Covid-19, significant correlations were found between apoptotic CEPs and RCP at enrollment, hemoglobin, MCH, MCV, platelets, and eosinophils. Apoptotic CECs correlated with RCP at enrollment and eosinophils.

**Fig.3:**
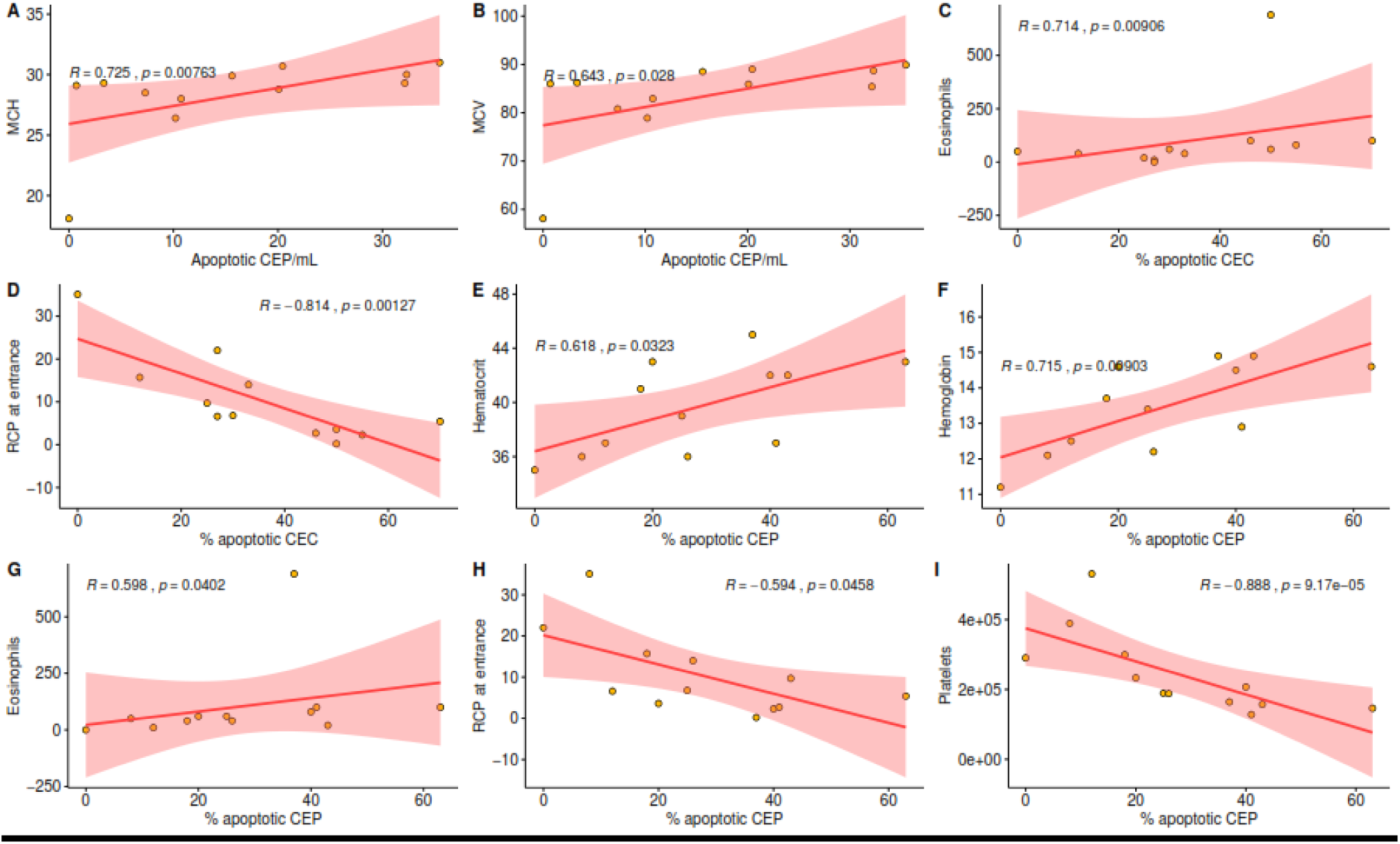
Significant correlations among CEP and CEP and hematological parameters for the in mild Covid-19 patients.

As shown in Fig. 4, in patients who recovered after Covid-19, and tested negative in the week before, significant correlations were found between CECs and red blood cell count, hemoglobin, hematocrit and MCH.

**Fig. 4:**
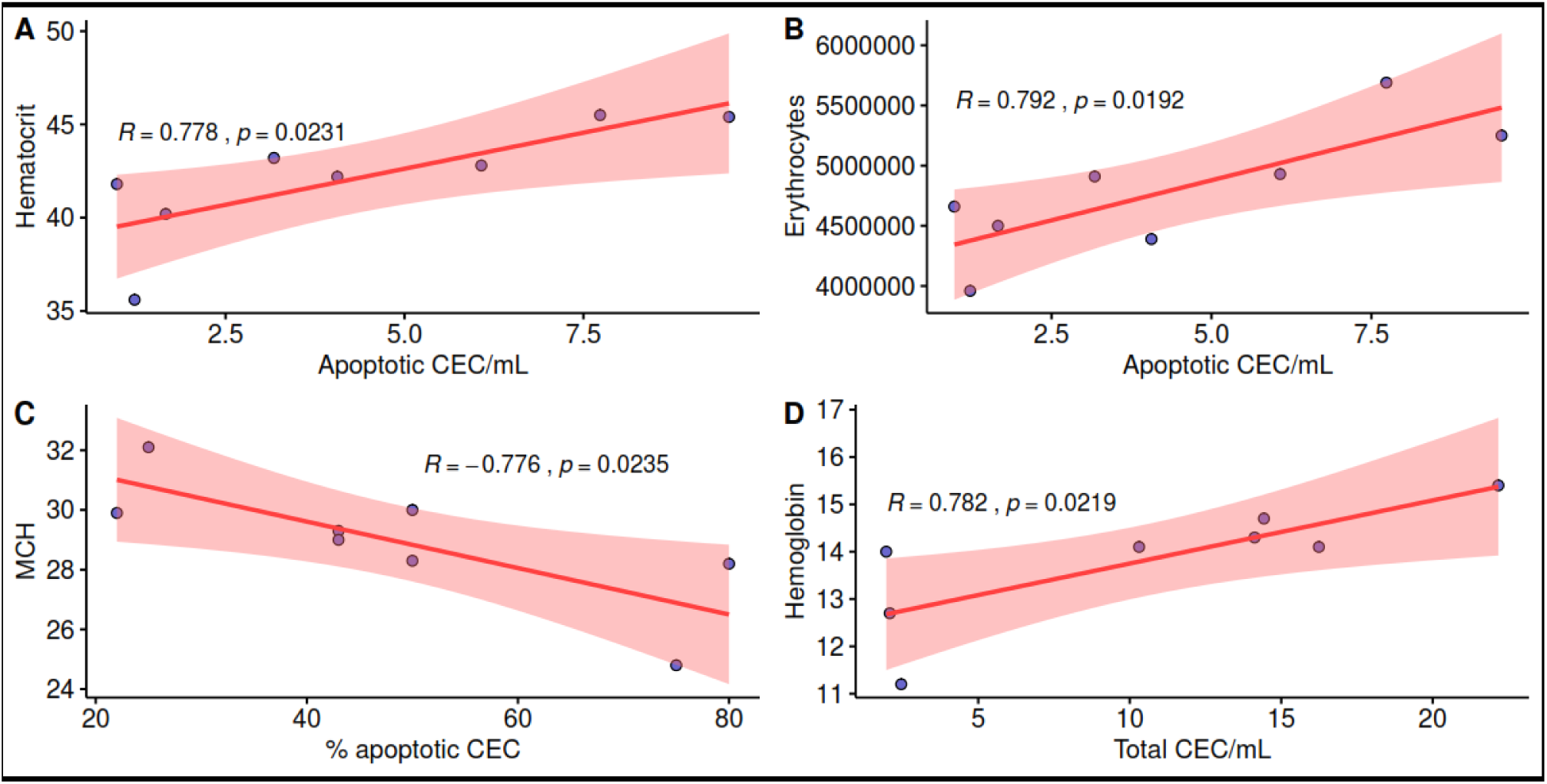
Significant correlations among CEP and CEP categories and hematological parameters for recovered patients.

## Discussion

There is emerging evidence that cardiovascular complications are frequently a crucial step in Covid-19 progression and related deaths (2–3). The SARS-CoV-2 virus infects patients by mean of the ACE2 receptor, which is widely expressed in organs in the respiratory tract, as well as in the kidney, intestine, and in the cardiovascular system. Endothelial cells express high levels of the ACE2 receptor, and a recent report has shown evidence of direct viral infection of endothelial cells and diffuse endothelial inflammation in Covid-19 patients (3). To our knowledge, this is the first report showing that CECs and CEPs are significantly unbalanced in Covid-19 patients with a pattern that was not previously reported in cancer, infectious and/or cardiovascular diseases.

Data reported here suggest that already in patients affected by mild Covid-19 disease CECs are significantly less apoptotic than in healthy controls, and that this unbalance is present also in severe Covid-19 patients. Both viable and apoptotic CEPs are significantly increased in mild and severe Covid-19 patients. Interestingly, Covid-19 patients who recovered from symptoms and tested negative in the week before had significantly less CECs when compared to controls and SARS-CoV-2-positive patients, whereas CEPs were still increased when compared to controls.

The mechanisms causing these unbalances deserve further translational investigation. The correlation found between copies of SARS-CoV-2 RNA and apoptotic CEPs, if confirmed, suggest the hypothesis that also these progenitors, in addition to mature endothelial cells might be a direct target of this pathogen. On the other hand, SARS-CoV-2-related endothelial damage might have triggered CEP mobilization from the bone marrow (or other reservoirs such as the white adipose tissue, see ref. 8) in an attempt to generate a newly functional vessel lining. These hypotheses should be investigated in adequate preclinical models.

Taken together, our findings indicate that CECs and CEPs deserve further study as possible candidate biomarkers of endothelial damage in Covid-19 patients, particularly in the early phases of the disease. This hypothesis is currently under investigation in our Institutions, and might pave the way to clinical trials using these biomarkers for patient selection and/or stratification.

## Data Availability

All raw data are available upon request

## Acknowledgments

Supported in part by AIRC and Italian Ministry of Health

